# *“Ich-Mupong”,* a swollen stomach: An ethnographic study of the daily lived experiences of schoolchildren in schistosomiasis high transmission areas along Lake Albert, Hoima District

**DOI:** 10.1101/2024.09.17.24313804

**Authors:** Odoi Paskari, Stella Neema, Fred Bateganya, Birgitte J. Vennervald, Shona Wilson

## Abstract

**Background:** Our primary focus was *Schistosoma mansoni* infection and schoolchildren. Within communities the social environment may promote individual risk of infection for the school-aged children. There will also be demographic groups who are not targeted or reached by preventive chemotherapy campaigns. The behaviours of these other groups will interact with those of school-aged children, resulting in further infection risk through exposure-related behaviours. Furthermore, perception of the disease may significantly influence the schoolchildren’s lived experience of the infection and associated disease. It is therefore crucial to document the daily experiences of schoolchildren living in schistosomiasis high transmission areas along Lake Albert, Hoima District.

**Methods:** An ethnographic study explored schoolchildren’s perspectives and daily life organisations that shape their risk of schistosomiaisis and their perceptions of the disease. The study was conducted between November 2022 and August 2023. It involved in-depth interviews with schoolchildren and their parents, key informant interviews, focus group discussions with schoolchildren, and participant observations. Data was analysed using a reflexive thematic analysis. Code reports were generated inductively using ATLAS.ti (Version 7).

**Results:** The study revealed a significant level of knowledge and awareness about schistosomiasis among schoolchildren. They had understanding of the risk factors, continued exposure, and experiences of illness, though they had little autonomy to address these through their own behaviour as they were influenced by the behaviour of others and macro-factors such as WASH provision and economic need. Study participants experienced individual-level effects of schistosomiasis such as educational impacts and isolation as a significant form of stigma.

**Conclusions:** There is a need for continued community sensitisation and awareness campaigns to address social stigma, educational impact, and contamination and exposure-related behaviours. National and regional policies and programmes on WASH, livelihood and poverty eradication programmes need to be revisited in schistosomiasis high transmission areas to help provide alternatives and improve schoolchildren’s lived experiences.

## Background

Schistosomiasis, also known as bilharzia, is a chronic disease caused by infection with blood flukes (trematode worms) of the genus *Schistosoma* (1). It is a neglected worldwide tropical disease (2) that disproportionately affects poor, vulnerable people who are unable to access health services routinely (1) and live without potable water and adequate sanitation (3). Globally, schistosomiasis causes significant morbidity (4) and affects over 111 million school-aged children (SAC) (5). In tropical and subtropical regions, especially in African countries, it is viewed as a significant public health problem (1, 6). The morbidity caused by schistosomes is not only immediate but has long-term effects on children’s health and development. SAC and preschool children, in particular, are the most vulnerable to overt disease (7, 8). They typically harbour the largest number of adult worms, with copious tissue-entrapped eggs causing systematic and organ-specific inflammation. Studies have shown that children can acquire schistosome infections within the first few months of life (9, 10), leading to initial organ damage and altered development in early life (4, 11), in addition to growth retardation, anemia, and neurocognitive deficits (4, 12, 13). The infection may cause physiological and developmental insults (4), and the severity of schistosomiasis’s impact on children’s health and development underscores the crucial need for early intervention and prevention.

In Uganda, most high infections are found amongst the shoreline communities of Lakes Albert, Victoria and Kyoga, and the Albert Nile (14). The disease is known to affect children and adults, but infection levels peak in 10 to 15-year-olds, our target age group (15, 16). Despite annual school-based delivery systems (preventative chemotherapy (PC) campaigns) since 2004 (17), the Lake Albert shoreline retains a high prevalence of infection among SAC (18). Stothard *et al.* reported that the levels of reinfection were so substantial that within six months of praziquantel administration, there was little concrete impact on local prevalence (7). Meta-analysis reveales that children are highly exposed to schistosomiasis due to their responsibility for water-related household chores and behaviour-related activities such as swimming and bathing in water containing the infective cercariae (19). A study along Lake Albert shoreline reported that school-aged children typically have up to 30 minutes of active daily water contact and additional covert passive water exposure (18). This critical social environmental aspect of schistosome transmission is framed by contamination and exposure-related behaviours (19–21). Our primary target, the schoolchildren, live in a community. Moreover, in this community, there are other groups, such as preschool children, out-of-school children and adults, who are not targeted or reached in by PC campaigns. Yet, schistosome eggs can be introduced to freshwater by any infected vertebrate host. After maturation in freshwater snail hosts (21), schistosome cercariae are often released in copious numbers, which have the potential to infect any demographic group, like SAC, due to exposure-related behaviours (22).

From that background, we sought to better understand schoolchildren’s daily lived experiences, focusing on their local understanding of schistosomiasis, the risk factors, signs and symptoms and associated stigma; providing greater insight into why transmission is high in the study area and the impact it has on the schoolchildren.

## Methods

### Study Area and Setting

The study was conducted in Kaiso and Buhirigi primary schools in Hoima district in the sub-counties of Kabaale and Kigorobya, along the shores of Lake Albert. Hoima District is located in the mid-western region of Uganda, with Hoima Municipality, the seat of the District Headquarters, around 200 km from Kampala. Hoima District stretches to the Lake Albert border with the Democratic Republic of Congo in the west.

### Study Design

An ethnographic study was designed to explore schoolchildren’s perspectives and organisation of their daily lives in schistosomiasis hotspots; exploring their perspective and gaining understanding of the world from their societal viewpoint (23). The 1^st^ author participated overtly in the daily lives of the study participants for an extended period of ten months from November 2022 to August 2023, listening to what was said, asking questions, and collecting whatever data were available to throw light on to the study. Participant observation, key informant interviews (KII), in-depth interviews (IDI) and focus group discussions (FGD) were employed within this long-term, holistic, and flexible approach to data collection (24, 25).

### Sample Size and Participant Selection

The study primarily involved pupils of Kaiso and Buhirigi Primary Schools, the participatory schools in the FibroScHot trial, in which this anthropology study is integrated. Most study participants speak Alur, and a few speak Runyoro, the two main local languages. Other participants included health workers, educators, parents/guardians, and Local Leaders. Participants were purposively selected (26, 27), and their inclusion was due to their experience and ability to provide quality information and valuable insight on the research topic (28). In addition their interest in the study, schistosomiasis, and health research was taken into consideration. All the study participants were identified with assistance from the FibroScHot Project Coordinators. Twenty IDIs with schoolchildren and 20 with parents were planned, with 19 schoolchildren and 19 parents IDIs successfully conducted. Eighteen of 20 planned key informant interviews (KIs) were conducted and 12 of 16 FGDs. The sample sizes were selected with data saturation in mind and the interview process terminated when staturation was reached (26).

### Data Collection

KIIs were conducted at the district, sub-county, and village levels. Participants included district officers, educators (head teachers and school teachers), health workers (FibroScHot coordinators, health assistants, VHTs and nurses), and community leaders.

The schoolchildren and their parents participating in IDIs were interviewed with the help of an in-depth interview guide. The interview was conducted at home and on weekends in the afternoon when children are home, as were their parents, and are done with household chores. All the interviews were conducted in Alur and translated into English with the help of an interpreter.

FGDs were conducted with purposively selected pupils from primary grades 4–7 who were 9– 17 years old. Each group consisted of six to ten participants. Group category included class and age. In total 86 schoolchildren participating in the FGDs. These interviews were conducted in Alur with the help of an interpreter.

Participant observations at school, home and in the community to document schoolchildren’s daily experiences in Kaiso and Buhirigi were also undertaken; overtly observing what happens, listening to what is said, and questioning participants (28). These observations were conducted over the whole 10-month data collection period.

### Data Management and Analysis

Data was analysed using reflexive thematic analysis following six steps as suggested by Braun and Clarke: Familiarisation, coding, generating initial themes, reviewing and developing themes, refining, defining and naming themes, and writing up (29). Familiarisation involved reading through the transcripts and taking down brief descriptions of potential codes and meanings. This was followed by inductive coding using ATLAS.ti (Version 7). ATLAS.ti was also used to generate coding reports. Each code developed was examined and what to merge or discard was determined. Initial themes were generated by the 1^st^ author grouping similar codes, followed by an iterative process of regrouping the themes with further review of the codes under each and their meanings. Theme development was a collaborative process, to achieve richer interpretations of meaning rather than attempting to achieve a consensus of meaning. Attention was paid to identifying the similarities and differences between participants’ views from the two study sites. All interviews from FGDs, KIIs, and IDIs were analysed to avoid missing essential data across the different participant groups. Evidence from across the identified themes is used to illustrate recurrent patterns that inform pupils’ understanding of three primary thematic areas: 1) understanding of illness, 2) the risk factors for bilharzia, and 3) the experience of bilharzia and its effects.

## Results

### Socio-Demographic Characteristics of the Participants

***IDI with schoolchildren*:** The IDI participants were 58% females and 58% Alur ethnicity, aged 10-16, with 63% being between 10-13 years. All reported knowledge of schistosomiasis (Supplementary Table 1).

***IDI with parents:*** The 19 adult participants were aged 44-62 years, were mostly fisher folks from Kaiso and agriculturists from Buhirigi, with a majority being married. Most had lower-level education, with only two having secondary education and one having a university degree (Supplementary Table 2).

**Focus Group Discussion (FGD) with Schoolchildren:** Six Focus Group discussions (FGDs) were conducted in Kaiso and Buhirigi Primary Schools respectively, mostly with single year groups drawn from primary 4 to 7 but 4 FGDs were of two consecutive year groups combined. Most participants were Alur-speaking, aged 10-17years, and reported having knowledge about schistosomiasis (Supplementary table 3).

**Key Informants Interviews (KIIs).** Out of 18 interviews, a quarter were with primary school teachers in government schools, over half were females and tertiary-level educated, and half were health workers at various levels (Supplementary Table 4).

### Understandings of illness

In this theme, the focus was on understanding the schoolchildren’s perceptions of illness in general and schistosomiasis in particular. Predominantly, the participants defined schistosomiasis by its associated signs and symptoms and viewed it as a water-borne disease without considering the context of the snail intermediate host. This was expressed by both pupils and key informants. The second most common definition provided by participants was exposure to water infested with snails. However, this was more common among the key informants than the children, with only one of the Buhirigi FGDs mentioning this.

> *It is a waterborne disease that may not be seen, but it stays in water. **(Female School child - IDI)***

When asked about the different ways pupils get schistosomiasis, they stated that fishing, bathing and playing in infested water, drinking infested water, and fetching water barefooted are among the ways pupils contract schistosomiasis. Others erroneously mentioned that one can get schistosomiasis by eating half-cooked fish and attending school barefoot. One pupil had this to say:

> *All I know is that if you bathe in the lake, the worms get to your body, and then I also know that when you defecate in the lake, the worms enter your body as you defecate also when you gas in the water, the worms enter your body after you have gassed. Also, if you are bathing in the lake and you accidentally drink the water, you get the disease. Other times, when it rains and the rains carry the stagnant water to the lake, and people fetch the water for cooking, they carry the worms in the water, and yet some of them even drink them. **(Female School child - IDI)***

We further asked the participants the local terminologies used to understand the illness. These largely depended on the participants’ ethnicity but there were also within ethnicity variations. Alurs from Congo called it *“bilharzo,”* as captured verbatim from one of the FGD participant: “For us from Congo, we are the ones who know it and call *bilharzo.”* Those from West Nile called it “*blazos*”, and as exemplified by a parent, “in Alur here, we mostly say schistosomiasis is a worm that disturbs children, but we also call it “*blazos*.” For the Banyoro, they called it “*empuuka*” meaning disease that makes stomach swell; the equivalent of which in Alur was “*ich-mupong*”. Others referred to it as “*koko*” meaning a worm. The local terminologies also shed light on children’s experience of schistosomiasis:

> *Even today, some children told me that somebody had that thing (“Koko” meaning a worm). The stomach started swelling. I said that one has schistosomiasis, and they said no, “Koko.” That worms. And that boy was thinking that if people would come, I would bring him, and they would see him because the boy started by feeling stomach pain like that, and his stomach had swollen. **(Health Worker - KII)***

## Risk factors for Schistosomiasis

In this theme we were interested in finding out the risk factors for schistosomiasis in the community. These were grouped according to micro and macro-level factors. The micro-level perspective looks at the individual manifestations of culturally prescribed behavioural patterns. Macro-level perspectives emphasise larger social forces as the ultimate risk factors for schistosomiasis, not the cumulative effects of individual effects *per se*.

## Mirco-level risk factors

### Poor hygiene and sanitation

Almost all participants mentioned poor hygiene and sanitation. When they talked about poor hygiene and sanitation, they referred to drinking untreated water, bathing in dirty water from the River/Lake, urinating and defecating in the lake or river, and poor disposal of faecal waste. In addition to defecating in water and open places, poor sanitation practices also included creating an outlet in latrines to allow free flow of faeces out when latrines become full. This practice was more common in Kaiso where it was common to come across faeces in open places and the foul smell of poorly disposed faecal waste. One FGD participant emphasised bathing in the lake and urinating and defecating in water as ways of spreading schistosomiasis.

> *Most children are getting the disease because when they are sent to the lake to wash clothes, they end up bathing in the lake, and while they bathe, some of them urinate and defecate in the water, and that is what makes them get the disease. **(Male School child - FGD)***

FGD participants of both sites reported legitimate beliefs and practices among the risk factors for schistosomiasis. Legitimate beliefs and practices include the belief that lake water quenches thirst, and the fear of falling in the latrine. One of the FGD participants stated that fear of falling in the latrine makes some children defecate outside, where other children step on it, resulting in transfer of faeces.

> *It is mostly the children who are in primary one that tend to get the disease because these children fear that they can fall inside the latrine, so they defecate outside of the latrine while the rest come and urinate on the faeces and others come and step on the faeces sometimes without even knowing. Those who step do not even wash their legs, and that is how they end up getting the disease. **(Male School child - FGD)***

### Choice of water source

Key informants and parents from Buhirigi reported that negative attitudes and perceptions towards borehole water among the risk factors. They said that water from the borehole is not sweet, it is salty, yet water from River Hoimo is not salty, so they would rather drink water from River Hoima. Others voiced that their parents and grandparents had been drinking water from River Hoimo, and nothing happened to them. So, there is no need for them to stop now.

> *I have a grandchild here with me who came from the lake shores. When this girl started staying with me here, she used to refuse to drink safe water from the borehole, claiming the water from the borehole was not sweet and, therefore, she preferred water collected from River Hoimo. So, she used to drink water from Rriver Hoimo until she started complaining of stomach pain. It is of recent that they tested her at the school for schistosomiasis, and she tested positive. They started her on the schistosomiasis medicine, and immediately, we started seeing some changes*. ***(Female Parent-IDI Buhirigi)***

### The cultural importance of swimming

Across the different groups and sites swimming, both as a sport and survival strategy, was acknowledged as a risk factors. Here, swimming is seen as a sport and an essential life skill, and adults spend time teaching children how to swim. Some pupils even escape school to go swimming, others swim during breaks and lunchtime. One of the key informants emphasised how important swimming is:

> *Swimming is a habit; it is like a sport for children. I would say it, ahh, for them, they take it as a sport. It is part of sports; it’s like when people go into the swimming pool to swim, it’s like part of the sport. So, these children, they go and take this with this part of the sport not by coincidence, but they love it because every time they are from school, they go swimming. **(District Officer - KII)***

One parent echoed the time pupils spend swimming:

> *Children play a lot in that water, most of them spend most of their time playing in that water, and they love playing in the water. When they go to shower, they spend the whole day swimming, and they come back home late. **(Female Parent - IDI Buhirigi)***

### Gender division of labour

Most of the community members including SAC heavily rely on the lake for their livelihood. This is so even when they know they may contract schistosomiasis from engaging in lake-based activities. One key informant equated the lake to a garden that requires daily visits.

> *If you tell someone not to go to the lake, then the person will ask you, where do you think our garden is? This is our garden. It is where we get money from. If we do not fish or…..go to prepare the net or the boat, where am I going to get the money? And then these young boys go there but for them every day in the lake. They will not go fishing but go there to prepare those things**. (Female Educator - KII)***

During the daily community participant observation along Lake Albert shoreline and River Hoimo, gendered lakeshore activities were observed. Both school-aged males and females were observed playing, walking and playing in the river/lake and associated swamps. Except for play, most of lakeshore activities had economic value with those participating paid for their services. In the morning between six and ten o’clock, SAC, especially boys, would be seen removing boats from the water and drying fishing nets. At the same time, the girls and their mothers wash fish and dry silverfish. In the afternoon, school-aged males would be seen setting up boats in preparation for evening fishing. Fishing with hooks continued throughout the day and night, with SAC of both sexes fishing during the day. A Key Informant and parent elaborate on the lakeshore activities:

> *The boat is pulled outside; they even enter the lake, which brings them into contact with the lake. When the workers come, when they reach the lake shore, they remove their luggage and leave the boat when it is still in the water but not very far, a place where someone can move and get that boat. So those children go and begin pushing the boats to bring it outside, so they enter into the water and not only that one maybe you find that some other bosses, let me call them bosses they give them the engine, and they tie it on the boat and move with that boat in the lake and go and leave the boats there. So, when they are coming back, they come back when they are swimming when they put on the life jacket. **(Male Health Worker - KII)***

> *It is because so many things happen on the shores; they wash fish from that same water that people step in as they go fishing. **(Female Parent - IDI Buhirigi)***

In Buhirigi, washing motorcycles by boys in River Hoimo was a routine activity.

> *Schistosomiasis is common because you find rivers like Hoimo people washing their motorcycles in it and also drinking it. **(Female School child - FGD)***

Participation in domestic chores were also among the lake shore and river-based activites. These domestic chores were performed and divided based on traditional gender roles and age, so conducted on the whole by females. These domestic chores included fetching lake/river water, washing utensils, and washing clothes in the lake.

## Macro-level risk factors for schistosomiasis

### Poor WASH infrastructure

Amongst macro-level risk factors reported were inadequate alternative water sources. The participants noted that water is scarce, with no boreholes and limited water sources. As a result of water scarcity, participants used water from the lake/river to meet their needs. Most of them see no other option but to collect and bathe in water from the lake/river. For instance, from one of the pupils:

> *It is because I bathe and drink water from river Hoimo, given the fact that that’s the only water we have, although sometimes I fetch the water and bring it home to first boil it before using it. **(Male School child - IDI)***

Schoolchildren and their parents in Kaiso and Buhirigi also stated that the wells, boreholes, and taps are far; thus, the long distance makes them resort to lake/river water to meet their needs. Even when participants wish to collect safe water, the distance to these water sources, if available, deters some, hence increasing cases of schistosomiasis in the community.

> *One may want to collect safe water for drinking, but when they imagine the distance, they immediately give up and decide to go and collect water from the river; after all, that is also water. **(Male School child - IDI)***

Closely related to the above was the existence of inadequate sanitation facilities. Both parents and pupils reported these views during in-depth interviews at both study sites, stating that toilets and latrines were few and some of them full, while some areas did not even have latrines. For instance, one of the pupils explained how inadequate sanitation facilities led to the spread of schistosomiasis.

> *Children also bathing in the lake can spread schistosomiasis because they bathe in the lake water. Some of them drink dirty water. Then, some other people in this community put pipes in their toilets to let out faeces when their latrines get full, and all these faeces end up going into the lake, and some children drink that lake water. **(Female School child - IDI)***

### Settlement patterns

Lastly, the study participants, especially those from Kaiso, noted the effect of settlement patterns on the spread of schistosomiasis. The settlement patterns result in some community members always being near the water. The dense concentration of people near Lake Albert’s lakeshore line, in violation of the recommended 400 meters, has increased the spread of schistosomiasis in these areas, as one of the key informants reported.

> *And of course, in terms of settlement, you realise, ahh, why the toilets are located up, it is because those soils up are better. Of course, we recommend it, but some issues need enforcement. The distance from the lake, together with the settlement of the people, the recommended distance is supposed to be around 400 meters, but you find people are just very near. This one actually creates a very big challenge because most of those families which are very close to the water source, mostly they are the ones who misuse that water source. So, you realise sanitation are still a very big challenge because you said you saw three public toilets. **(District Officer - KII)***

## The experience of Schistosomiasis

In this theme, we aimed to explore the perceptions of the prevalence of schistosomiasis in the community and the resulting physical and social impacts. Specifically, we sought to understand the signs and symptoms of the disease and whether they are considered serious or something individuals simply learn to cope with. Additionally, we aimed to uncover any social stigma associated with these symptoms and how individuals respond to their experiences with the disease. During the study, all participants acknowledged that schistosomiasis is prevalent in the two study sites. When asked about personal experiences with the disease, the majority confirmed having had it. When asked to define it, the participants most frequently defined schistosomiasis based on its signs and symptoms, which included bloody and non-bloody diarrhoea, and swollen stomach. This was especially prevalent amongst those from Kaiso Primary, and was identified through individual interviews and focus group discussions.

> *What I know about schistosomiasis is that they start as worms, and these worms burn your stomach and cause diarrhoea. The worms also make your stomach swell. So, if someone portrays the signs that I have told you, that means that the person is suffering from schistosomiasis. **(Female School child - IDI)***

Other symptoms described included loss of appetite and energy, lower abdominal pain, itching, persistent sickness, headache, changes in urine and skin colour, vomiting blood and for some a sense of impending death or loss of bodily senses.

> *I feel my stomach is paining, and I feel like I want to sleep, then I sleep, but when I wake up, I feel like not eating that is all. **(Female School child - IDI)***

> *I had terrible headaches, and I also suffered from stomach pain; I would also vomit and feel a lot of pain. **(Female School child - IDI)***

The most widely reported outwardly recognisable sign was swollen body parts. This was broken down into “*ichapongaponga*”, meaning swollen stomach, big and hard stomach, big but empty stomach or big stomach in Alur, but also mentioned were swollen cheeks, swollen legs and swollen feet.

> *There are many children suffering from it. There is some boy up there; his stomach is big, and he over bathes in the lake. **(Male School child - IDI)***

As a result of the above signs and symptoms, participants from both sites reported social stigma associated with schistosomiasis. This manifested in isolation, verbal abuse, criticism, labelling, being beaten and laughed at. The most commonly reported social stigma was isolation. Participants reported that when someone was suspected of having schistosomiasis, he/she would be chased by others, avoided and not played with, not invited to participate in games that were ongoing, and games would end if they were around.

> *Sometimes, they know that you have the disease, so they think that when we go and sit with that person, I may get infected; it is better such a person plays alone, and I also play alone with those without the disease. **(Male School child - FGD)***

This isolation was echoed by a Key Informant:

> *They feel so bad because they see them not feeling well with their swollen stomach and big feet, and these people who are okay always laugh at those who are sick, thereby starting to isolate them when they no longer socialise with them. **(Male Health Worker - KII)***

Study participants also reported educational effects – loss of concentration and poor performance in class, and interruptions in learning through missed lessons, school absenteeism, and dropping out of school. For example, one of the FGD participants mentioned being unable to go to school because of having schistosomiasis.

> *I feel bad in that case because I am unable to go to school yet my friends are going to school. **(Female School child - FGD)***

One of the key informants reiterated the effect on children’s education.

> *At the individual level, like our children, it affects their learning because if a child is having, maybe is not feeling well, it will make the child be absent. At times, it even leads to school dropout. If the child is not feeling well, the child says, ahh, I have left schooling, so sometimes it leads to school dropout. **(Male Educator - KII)***

## Discussion

Research shows that young people have unique ideas about illness, its causes and cure, which is influenced by media, education, family, and personal experiences. These views often mirror adults’ but sometimes differ significantly (23). Some recent studies (30–32) have pointed out that younger children generally have low and variable levels of knowledge about both disease and interventions. However, in the participants understood schistosomiasis through its signs and symptoms, its health impact, and its water-borne nature, underscoring the schoolchildren’s agency as actors who are knowledgeable about the disease and can express their voices.

Any medical anthropological study that hopes to shed light on the disease behaviour connection must ultimately adopt a micro-macro perspective. The macro-risk of lack of or inadequate sanitation facilities most especially along the shoreline communities, such as those observed in Kaiso has made schistosomiasis a challenging disease to control in sub-Saharan Africa (14). However, Rollinson (33) warns that adequate sanitation facilities do not necessarily guarantee its use. In line with other studies (34–36), poor hygiene and sanitation related behaviours were identified as a micro-risk behaviours, most especially in Kaiso. Therefore, even when latrines are constructed and existing ones improved, health education and sensitisation ought to be integrated. At the macro level, an argument is made that increasing access to safe water is an intervention that will significantly reduce the risk of schistosomiasis transmission (22, 36). Our study found both through interviews and observations that inadequate alternative water sources and the long distance to the available safe water sources were risk factors for schistosomiasis. The World Health Organization calls for the provision of safe water, sanitation, and hygiene as one of the five key interventions within the global NTD roadmap (37). Our study shows we still have a long way to go to address safe water issues in Ugandan lakeshore communities.

Equally important to WASH related risks were the roles played by lake shore activities in the spread of schistosomiasis. These lakeshore activities had economic value. Other studies in Kenya (38, 39) have equally reported that the pursuit of income-generating activities such as fishing may predisposed participants to infection. Such participation could also be attributed to differences in childhood experiences. According to Helman (23), in poorer societies, children are, in effect, ‘trainee adults’, expected to perform almost all the usual adult tasks, such as childcare, cooking, hunting, herding and earning money, as early as possible; and warned that in understanding health and illness, it is important to avoid seeing the poor health of a population as the sole result of its culture without looking at its particular economic or social situation. A case in point, it was observed that there was a heavy reliance on the lake for economic needs in Kaiso by both children and adults. SAC, particularly boys, would be seen going to the lake everyday to bring boats from the water, remove nets from the boats, dry the nets and prepare fishing gear and they were paid for their services. Similarly, in Buhirigi SAC would be seen washing motorcycles for a pay. Such results reveal not only the cultural perspective but also the economic and social situation of the participants in the two sites. As a result, there will be need for mindset change and livelihood diversification since the family social structure allows SAC to participate in livelihood related activties. It is also important that domestic activities were undertaken along the lakeshore particularly by girls. A linkage between performance of domestic chores and schistosomiasis is well established (40, 41). Gender therefore played a significant role in the types of micro-risk behaviours observed but all age groups are at risk through different water contact activities as reported elsewhere (18). Participants acknowledged that schistosomiasis is common in the two study sites attributed this to the reported and observed frequent water contact, in line with previous findings in Ugandan lake shore settings (18).

The most widely reported signs and symptoms by the participants were swollen body parts. Although through observation these signs and symptoms was not as readily pronounced as the results of the interviews suggest, a few SAC with symptoms were seen. Schistosomiasis has previously been suggested to carry with it a social burden of stigma (42), as reported for both *S. mansoni* (43) and *S. haematobium* endemic areas (43, 44) despite the differing manifestations of the infections. Capturing of “stigma” as nuanced in English can be influenced by how easily the concept can be translated to other languages (43). A strength of our study is therefore the capturing of “stigma” through assessment of lived experience utilising a range of qualitative methods. Participants reported that as a result of suffering from schistosomiasis, they experienced isolation, verbal abuse, criticism, labelling, beating, and laughing, whilst during community and school visits, isolation and labelling as forms of stigmatisation of children, most especially those with swollen stomachs was observed. Triangulation of the methods, therefore, indicates that stigmatisation of children suffering from schistosomiasis in these communities is a significant reality, and more can be done to address this directly in health education campaigns.

Finally, in line with others, (45, 46) our study found educational impacts. This reduced educational performance and school attendance can impact future adult productivity, and wage-earning capacity (47, 48). To negate this negative impact on the academic and future productivity continued sensitisation promoting adherence to treatment programmes is required.

## Study limitations

We stress that our study does not aim at generalizing our findings but rather to provide indepth analysis of schoolchildren’s lived experiences in the two subcounties. Generalization, will only be achieved through replication in diverse communities. Furthermore, our focus was on school-going childen, not out-of-school children, adults and preschoolers and yet their lived experiences could provide additional vital lessons.

## Conclusions

It is evident that there is a high level of knowledge and awareness on schistosomiasis, including what it is, its mode of transmission, risk factors, signs and symptoms and resultant stigma. The micro-level risk factors featured more prominently than macro-level level factors, though they were significantly inter-linked. Therefore, attention must be paid to addressing contamination and exposure-related behaviours since most risk factors rotate around this continuum. The lack of alternative livelihoods calls for livelihood diversification and regional-specific poverty eradication programmes. Study participants experienced learning impacts and isolation as a significant form of stigma as a result of schistosomiasis. Thus, community sensitisation and awareness campaigns need to be integrated with continued preventive chemotherapy to reverse this trend.

## Data Availability

The Makerere University School of Social Sciences Research Ethics Committee-approved study Participant Information Sheets do not permit sharing of the original transcripts. The transcript excerpts that support the conclusions of this article are included within the article and its additional files

## Acknowledgements

When drafting this manuscript, the first author, Mr. Odoi Paskari, was a postgraduate student in Medical Anthropology at the Department of Sociology and Anthropology, School of Social Sciences, Makerere University. We are grateful for the support from the Department and the implementation research team under the FibroScHot project. The authors would also like to acknowledge the contribution of the school children, parents/guardians, teachers and administrators from Kaiso and Buhirigi primary schools, Hoima District Health Office and the community members of Kabaale, Buseruka and Kigorobya Sub counties, who were very supportive and ensured that this study was conducted without any significant challenges. We also acknowledge contributions from Jovia Adubango, the graduate student who worked alongside the 1^st^ author as an interpreter during data collection.

## Ethical approval, consent and assent to participants

The study was approved by the Makerere University School of Social Sciences Research Ethics Committee (MAKSSREC REF 08.2022) and the Uganda National Council for Science and Technology (UNCST REF No SS1463ES). Still, we received clearance from Hoima district, school authorities of Kaiso and Buhirigi, and community leaders before conducting the study. Written informed consent and assent were obtained from study participants using the ethical consent and assent process, and confidentiality of information and privacy was ensured by concealing participants’ identities, conducting sessions in private places, and refraining from asking for information that would compromise participants’ privacy.

## Authors’ contributions

OP: Writing – review & editing, Writing – original draft, Methodology, Data curation, Formal analysis, Conceptualisation.

SN: Writing – review & editing, Funding acquisition, Project administration, Supervision

FB: Writing – review & editing, Supervision

BV: Writing – review & editing, Funding acquisition,

SW: Writing – review & editing, Funding acquisition, project administration

