## Supplementary tables 1 to 4 for "*“Ich-Mupong”,* a swollen stomach: An ethnographic study of the daily lived experiences of schoolchildren in schistosomiasis high transmission areas along Lake Albert, Hoima District"

**Supplementary Table 1: Socio-demographic characteristics of the in-depth interview**

**Participants (Schoolchildren)**

| **Gender** | **N (%)** |
|---|---|
| Male | 8 (42%) |
| Female | 11 (58%) |
| **Age range** | |
| 10-13 | 12 (63%) |
| 14-17 | 7 (37%) |
| **Class** | |
| Primary seven | 2 (11%) |
| Primary six | 4 (21%) |
| Primary five | 7 (37%) |
| Primary four | 6 (31%) |
| **Ethnicity** | |
| Alur | 11 (58%) |
| Mugungu | 3 (16%) |
| Munyoro | 2 (10%) |
| Others (Lugbara, Jukoth, Congolese) | 3 (16%) |
| **School** | |
| Kaiso Primary School | 10 (53%) |
| Buhirigi Primary School | 9 (47%) |
| **Heard of Bilharzia** | |
| All | 19 (100%) |

**Supplementary Table 2: Socio-demographic characteristics of the in-depth interview participants (Parents)**

| **Gender** | **N (%)** |
|---|---|
| Male | 7 (37%) |
| Female | 12 (63%) |
| **Age range** | |
| 27-37 | 5 (26%) |
| 38-47 | 7 (37%) |
| 48-57 | 5 (26%) |
| 58-67 | 2 (11%) |
| **Education** | |
| None | 2 (11%) |
| Primary | 13 (68%) |
| Secondary | 3 (16%) |
| Tertiary | 1 (5%) |
| **Marital status** | |
| Married | 17 (89%) |
| Widow | 2 (11%) |
| Occupation | |
| Farmers | 7 (37%) |
| Fishing business | 8 (42%) |
| Fisherman | 2 (11%) |
| Other | 2 (10%) |
| **Ethnicity** | |
| Alur | 14 (74%) |
| Others (Munyoro, Mugungu and Japadhola) | 5 (26%) |
| **Site** | |
| Kaiso primary school | 10 (53%) |
| Buhirigi primary school | 9 (47%) |

**Supplementary Table 3: Socio-demographic characteristics of the Focus group discussion**

**participants (schoolchildren)**

| FGD | School | Number of participants | Age range | class | Gender | Heard of Schistosomiasis |
|---|---|---|---|---|---|---|
| 1 | Buhirigi | 7 | 14-17 | Primary Seven | Male & Female | Yes |
| 2 | Buhirigi | 7 | 13-16 | Primary Six | Male & Female | Yes |
| 3 | Buhirigi | 8 | 12-16 | Primary Seven and Six | Male & Female | Yes |
| 4 | Buhirigi | 7 | 12-14 | Primary Five | Male & Female | Yes |
| 5 | Buhirigi | 10 | 12-15 | Primary Four | Male & Female | Yes |
| 6 | Buhirigi | 8 | 10-15 | Primary Five and Four | Male & Female | Yes |
| 7 | Kaiso | 6 | 14-16 | Primary Seven | Female | Yes |
| 8 | Kaiso | 6 | 14-16 | Primary Six | Male & Female | Yes |
| 9 | Kaiso | 6 | 11-15 | Primary Seven and Six | Female | Yes |
| 10 | Kaiso | 6 | 12-14 | Primary Five | Male & Female | Yes |
| 11 | Kaiso | 7 | 11-13 | Primary Four | Male & Female | Yes |
| 12 | Kaiso | 8 | 10-13 | Primary Five and Four | Male & Female | Yes |

**Supplementary Table 4: Socio-demographic characteristics of the key informants**

| **Gender** | **N (%)** |
|---|---|
| Male | 8 (44%) |
| Female | 10 (56%) |
| **Level** | |
| School | 8 (44%) |
| Community | 6 (33%) |
| Sub-county and District | 4 (22%) |
| **Designation** | |
| Health worker | 9 (50%) |
| Educator | 6 (33%) |
| Government | 3 (17%) |
| **Education** | |
| Secondary | 12 (66%) |
| Diploma | 3 (17%) |
| Degree | 2 (11%) |
| None | 1 (6%) |
